# SARS-CoV-2 antibody responses in children with MIS-C and mild and severe COVID-19

**DOI:** 10.1101/2020.08.17.20176552

**Authors:** Elizabeth M. Anderson, Caroline Diorio, Eileen C. Goodwin, Kevin O. McNerney, Madison E. Weirick, Sigrid Gouma, Marcus J. Bolton, Claudia P. Arevalo, Julie Chase, Philip Hicks, Tomaz B. Manzoni, Amy E. Baxter, Kurt P. Andrea, Chakkapong Burudpakdee, Jessica H. Lee, Laura A. Vella, Sarah E. Henrickson, Rebecca M. Harris, E. John Wherry, Paul Bates, Hamid Bassiri, Edward M. Behrens, David T. Teachey, Scott E. Hensley

**Affiliations:** Department of Microbiology, Perelman School of Medicine, University of Pennsylvania, Philadelphia, PA USA; Immune Dysregulation Frontier Program, Department of Pediatrics, Children’s Hospital of Philadelphia, University of Pennsylvania Perelman School of Medicine, Philadelphia, PA, USA; Division of Oncology, Department of Pediatrics, Children’s Hospital of Philadelphia, University of Pennsylvania Perelman School of Medicine, Philadelphia, PA, USA; Division of Rheumatology, Department of Pediatrics, Children’s Hospital of Philadelphia, University of Pennsylvania Perelman School of Medicine, Philadelphia, PA, USA; School of Veterinary Medicine, University of Pennsylvania, Philadelphia, PA USA; Institute for Immunology, Perelman School of Medicine, University of Pennsylvania, Philadelphia, PA, USA; Department of Systems Pharmacology and Translational Therapeutics, University of Pennsylvania, Philadelphia, PA, USA; Division of Infectious Diseases, Department of Pediatrics, Children’s Hospital of Philadelphia, University of Pennsylvania Perelman School of Medicine, Philadelphia, PA, USA; Division of Allergy and Immunology, Department of Pediatrics, Children’s Hospital of Philadelphia, University of Pennsylvania Perelman School of Medicine, Philadelphia, PA, USA; Department of Pathology and Laboratory Medicine, Children’s Hospital of Philadelphia, University of Pennsylvania Perelman School of Medicine, Philadelphia, PA, USA; Penn Center for Research on Coronavirus and Other Emerging Pathogens, University of Pennsylvania Perelman School of Medicine, Philadelphia, PA, USA

**Keywords:** COVID-19, Pediatric, Antibodies, Multisystem Inflammatory Syndrome in Children (MIS-C), SARS-COV-2

## Abstract

SARS-CoV-2 antibody responses in children remain poorly characterized. Here, we show that pediatric patients with multisystem inflammatory syndrome in children (MIS-C) possess higher SARS-CoV-2 spike IgG titers compared to those with severe coronavirus disease 2019 (COVID-19), likely reflecting a longer time since onset of infection in MIS-C patients.

## MAIN TEXT

Severe acute respiratory syndrome coronavirus 2 (SARS-CoV-2) manifests differently in pediatric populations. While the absolute numbers and rate of development of severe coronavirus disease 2019 (COVID-19) is significantly lower in children compared to adults (1), some pediatric patients develop severe to critical illness. Unlike adults, pediatric patients can also become afflicted with multisystem inflammatory syndrome in children (MIS-C) (2, 3). MIS-C is a syndrome that affects previously healthy children and manifests as a hyperinflammatory syndrome with multiorgan involvement that has some overlapping clinical features with Kawasaki disease shock syndrome (4–9). While it is believed that MIS-C represents a post-infectious sequela of SARS-CoV-2, the pathophysiology of this syndrome has not yet been delineated.

We sought to determine the humoral responses to SARS-CoV-2 in children presenting with COVID-19 vs. MIS-C to help illuminate potential pathophysiologies induced by the virus. We analyzed serum samples from 29 SARS-CoV-2 infected children admitted to the Children’s Hospital of Philadelphia (CHOP) in April and May 2020 (Supplementary table S1). We categorized these patients into three clinical disease phenotypes: minimal COVID-19 (asymptomatic children, or those with minimal symptoms; n = 10), severe COVID-19 (children requiring invasive respiratory support or an increase in positive pressure ventilation above their baseline; n = 9), and those with MIS-C (children meeting Centers for Disease Control criteria (2, 10, 11); n = 10). Detailed case studies of 6 of the 10 children with MIS-C (CD12, CD18, CD19, CD22, CD24, and CD26) were previously reported by our group (7). As expected, cycle threshold (Ct) values of SARS-CoV-2 RT-PCR were significantly lower in the pediatric patients presenting with severe COVID-19 (median: 28, IQR: 26 – 29) compared to children with MIS-C (p = 0.002 in one-way ANOVA) (Supplementary table S1). Similar to other reports (3), we found that children with SARS-CoV-2 had systemic inflammation evidenced by elevated inflammatory markers, including ESR, CRP, ferritin, and D-dimer (10). The severe COVID-19 and MIS-C patients also displayed elevated pro- and anti-inflammatory plasma cytokines (4, 5, 8). Finally, B-type natriuretic protein (BNP), a marker of cardiac inflammation, was higher in the MIS-C group versus the severe COVID-19 group, with the difference approaching statistical significance (Supplementary table S1).

We performed ELISAs to measure serum IgG antibodies against the SARS-CoV-2 full-length spike protein (S), the receptor binding domain (S-RBD) of the S protein (12, 13), and the nucleocapsid (N) protein (Figure 1A-C). We found that children in the minimal COVID-19 cohort had varied levels of serum IgG against all SARS-CoV-2 antigens tests (Figure 1), which likely reflects the clinical heterogeneity of these patients. These patients were either completely asymptomatic with respect to SARS-CoV-2 (n = 2), or were admitted for treatment of another infection (n = 3). In contrast, we found that the majority of children with severe COVID-19 had undetectable levels of SARS-CoV-2 S, S-RBD, and N IgG antibodies (Figure 1A-C). This observation stands in contrast to that in adults with severe COVID-19, who typically possess higher levels of SARS-CoV-2 antibodies compared to adults with milder disease (14, 15). We found that patients with MIS-C had higher IgG antibody titers against S-RBD and full-length S (p = 0.010 and p = 0.025 in one-way ANOVA, respectively) compared to children with severe COVID-19 (Figure 1C). Children with MIS-C also had elevated levels of serum anti-SARS-CoV-2 N antibodies; however, this was not significantly higher than children with minimal or severe disease. We also performed ELISAs to measure serum IgM and IgA antibodies against the SARS-CoV-2 S, S-RBD, and N proteins (Figure 1D-I). Unlike IgG titers, we found no statistically significant differences in IgM antibody titers between children with different SARS-CoV-2 diseases. We found that children with MIS-C had higher IgA antibody titers compared to children with severe COVID-19 against full-length S but not S-RBD (p = 0.010 in one-way ANOVA).

**Figure 1.**
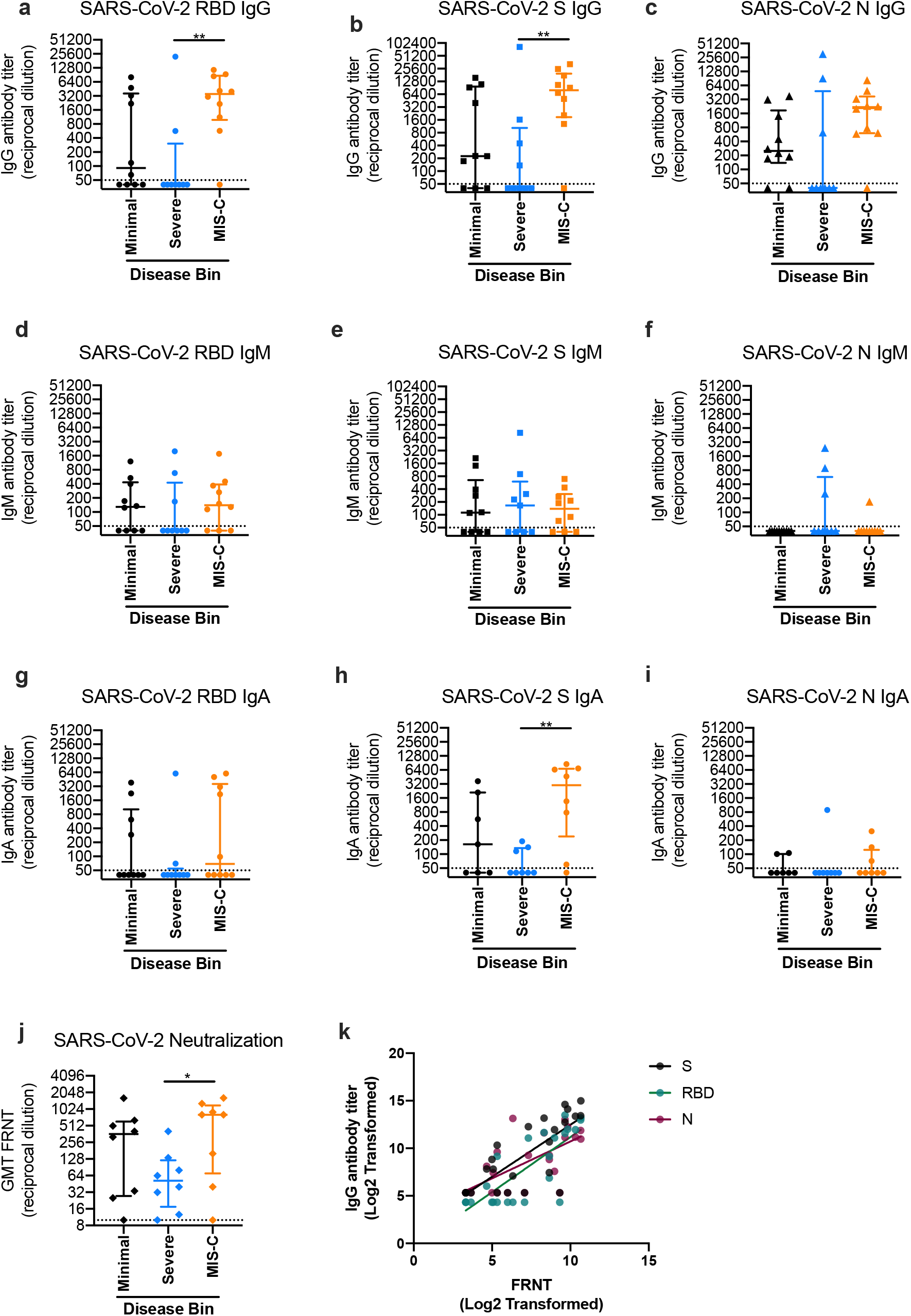
Serum SARS-CoV-2 antibody levels in pediatric COVID-19 patients. Antibody titers expressed as reciprocal serum dilution against SARS-CoV-2 antigens in pediatric patients with minimal disease (n = 10), severe disease (n = 9) and multisystem inflammatory syndrome (MIS-C; n = 10). Line and error bars represent median antibody titer and interquartile range per disease phenotype. Titers against the SARS-CoV-2 receptor binding domain (S-RBD) IgG (a), IgM (d), and IgA (g). Titers against SARS-CoV-2 full length spike protein (S) IgG (b), IgM (e), and IgA (h). Titers against SARS-CoV-2 nucleocapsid protein (N) IgG (c) and IgM (f) and IgA (i). Note: IgA S and N antibodies were measured in a subset of samples with sufficient volume; N = 23). (j) Neutralization activity of sera against SARS-CoV-2 spike pseudo-typed vesicular stomatitis virus (VSV) expressed as the geometric mean of the reciprocal dilution foci reduction neutralization titer (GMT FRNT; N = 24). (k) Linear regressions of Log2 transformed SARSCoV-2 IgG titers (S, S-RBD, and N) and FRNT neutralization titers. Dashed lines denote the lower limit of detection at a reciprocal dilution of 50. Unpaired t-test of log2 transformed titers

To measure levels of functional antibodies in pediatric patients, we also performed neutralization assays using pseudo-typed vesicular stomatitis virus (VSV) expressing the SARS-CoV-2 S protein (Figure 1J). Neutralization antibody titers highly correlated with IgG titers to full length S, S-RBD, and N (R^2^ = 0.586, 0.632, and 0.4643, respectively; Figure 1K). We found that children who presented with minimal disease had variable levels of neutralizing SARS-CoV-2 antibodies (Figure 1J). Children with MIS-C had higher neutralization titers compared to children with severe COVID-19 (Figure 1J), which is consistent with higher serum IgG titers against full length S (Figure 1A) and S-RBD (Figure 1B) in MIS-C children.

Collectively, our study suggests that children with MIS-C have higher levels of IgG antibodies that neutralize SARS-CoV-2 more effectively compared to children with severe COVID-19. Although this observation will require further study, we suspect that this finding may be due to a longer time since onset of infection in children with MIS-C relative to children with severe COVID-19. We could not formally investigate this possibility, since many of the patients in the MIS-C cohort did not recall a specific exposure or disease symptoms. Our previous studies indicate that adults with severe COVID-19 possess higher titers of SARS-CoV-2 S-RBD antibodies compared to adults with milder disease (14, 15). It is interesting that only 2 of 9 pediatric patients with severe COVID-19 had detectable IgG antibody titers against the S-RBD protein. One of these seropositive patients presented with severe COVID-19 associated acute respiratory distress syndrome (ARDS) in the setting of pre-existing hypertension, insulin-dependent diabetes mellitus and hypertrophic cardiomyopathy and eventually died from cardiac causes (Diorio et al, 2020 in press). The other seropositive patient had a history of adrenal insufficiency due to panhypopituitarism and presented with hypotension leading to respiratory failure, in the setting of multiple co-infections including rhinovirus, adenovirus, and a radiologically confirmed osteomyelitis. Further studies are required to determine why children with severe COVID-19 tend to have lower titers of SARS-CoV-2 antibodies compared to adults with similar disease.

## METHODS

### Study participants

We enrolled patients based on evidence of past or active SARS-CoV-2 infection (by positive RT-PCR in blood, stool or mucosa, the presence of serum IgG to SARS-CoV-2) or a very high clinical suspicion of MIS-C (10). Patients were categorized into three diseases phenotypes (MIS-C, severe COVID-19, or minimal COVID-19) after enrollment into the study. Patients were categorized as having MIS-C per the CDC case definition of MIS-C (11). Patients who presented with a primarily respiratory process requiring an increase in positive pressure support above their baseline and did not meet the criteria for MIS-C were categorized as “severe COVID-19”. Patients were classified as “minimal COVID-19” if they required hospitalization but did not otherwise meet criteria for MIS-C or severe COVID-19. Co-infections were identified by chart review for microbiologically proven infections that were deemed clinically significant by a panel of infectious disease physicians. This study was approved by the institutional review board at the Children’s Hospital of Philadelphia. Verbal informed consent was obtained from patients or their guardians in accordance with the Declaration of Helsinki. Due to the COVID-19 pandemic, verbal consent was obtained and written consent was signed by the consenting physician. All participants were provided with a paper copy of the consent form.

### Detection of SARS-CoV-2 Nucleic Acid

A real time-PCR assay for SARS-CoV-2 RNA was performed in a CLIA certified high-complexity clinical laboratory using a laboratory developed test with emergency use authorization from the FDA. The assay contained a primer/probe set for amplification and detection of the N2 gene of SARS-CoV-2 multiplexed with a primer/probe set for amplification of human b-actin as an internal control. RNA extraction from clinical samples was performed using the Roche MagNA Pure LC Total Nucleic Acid automated extraction platform. RT-PCR was performed using the Applied Biosystems Quant Studio DX using TaqMan chemistry. In this method, if a target is present, an increase in fluorescence during thermocycling is detected due to DNA polymerase cleavage of a TaqMan probe when the probe is bound, separating reporter and quencher dyes. A test is positive if measured fluorescence crosses a defined threshold above background levels, and the cycle threshold (Ct) is the number of cycles required for this to occur. Generally, the greater the amount of target nucleic acid present in the sample, the lower the Ct. A Ct of 45 or lower for the SARS-CoV-2 N2 target was defined as a positive result.

### Quantification of SARS-CoV-2 serum antibody titers

Serum IgG, IgM, and IgA antibody titers against SARS-CoV-2 antigens were quantified by enzyme-linked immunosorbent assays (ELISA) as previously described (13). Plasmids encoding the full-length SARS-CoV-2 spike (S) protein and the receptor binding domain (S-RBD) were provided by Florian Krammer (Icahn School of Medicine at Mt. Sinai, New York City NY).

SARS-CoV-2 S-RBD and the full-length S proteins were purified from 293F transfected cells by Ni-NTA resin. SARS-CoV-2 nucleoprotein (N) was purchased (Sino Biological; Chesterbrook PA) and reconstituted in Dulbecco’s phosphate buffered saline (DPBS). In brief, 200 μL of blocking buffer (DPBS supplemented with 3% milk and 0.1% Tween-20) was added to ELISA plates (Immulon 4 HBX, Thermo Fisher Scientific, Waltham MA) that were washed three times with PBS plus 2% Tween (PBS-T) after being coated overnight at 4°C with 2 μg/mL SARS-CoV-2 antigens. Sera were heat-inactivated prior to serial dilutions starting at 1:50 in dilution buffer (DPBS supplemented with 1% milk and 0.1% Tween-20). After blocking, ELISA plates were washed 3 times with PBS-T and 50 μL of diluted sera was added. After 2 hours of incubation, ELISA plates were washed 3 times with PBS-T and 50μL of secondary antigen at 1:5000 dilution (IgG; Jackson ImmunoResearch Laboratories; West Grove PA), 1:1000 (IgM; SouthernBiotech; Birmingham AL), or 1:500 (IgA; SouthernBiotech; Birmingham AL) in dilution buffer was added. ELISA plates were incubated for 1 hour, washed again 3 times with 300μL PBS-T, and developed by adding 50 μL of SureBlue tetramethylbenzidine substrate (SeraCare; Milford MA) and stopping the reaction with 25 μL of 250 mM HCl after 5 minutes. Optical densities at 450 nm wavelength were obtained on a SpectraMax 190 microplate reader (Molecular Devices, San Jose CA). Serum antibody titers were expressed as the reciprocal serum dilution at a set OD that was based off of a standard curve from the monoclonal antibody CR3022 starting at 0.5 μg/mL (for RBD and S ELISAs) or serially diluted pooled serum from actively SARS-CoV-2 infected adults (for N ELISAs). The plasmids to express CR3022 were a provided by Ian Wilson (Scripps Research Institute, San Diego CA). Standard curves were included on every plate to control for plate-to-plate variation.

### Production of VSV pseudotypes with SARS-CoV-2 S for neutralization assays

293T cells plated 24 hours previously at 5 × 10^6^ cells per 10 cm dish were transfected using calcium phosphate with 35μg of pCG1 SARS-CoV S delta18 expression plasmid encoding a codon optimized SARS-CoV S gene with an 18 residue truncation in the cytoplasmic tail (kindly provided by Stefan Pohlmann (German Primate Center, Göttingen, DE). 12 hours post transfection the cells were fed with fresh media containing 5mM sodium butyrate to increase expression of the transfected DNA. 30 hours after transfection, the SARS-CoV-2 spike expressing cells were infected for 2-4 hours with VSV-G pseudo-typed VSVΔG-RFP at an MOI of ∼1-3. After infection, the cells were washed twice with media to remove unbound virus. Media containing the VSVΔG-RFP SARS-CoV-2 pseudo-types was harvested 28-30 hours after infection and clarified by centrifugation twice at 6000g then aliquoted and stored at −80°C until used for antibody neutralization analysis.

### Antibody neutralization assay using VSVΔG-RFP SARS-CoV-2

All sera were heat-inactivated for 1 hour at 55°C prior to use in neutralization assay. Vero E6 cells stably expressing TMPRSS2 were seeded in 100 μl at 2.5×10^4^ cells/well in a 96 well collagen coated plate. The next day, 2-fold serially diluted serum samples were mixed with VSVΔG-RFP SARS-CoV-2 pseudo-type virus (50-200 focus forming units/well) and incubated for 1hr at 37°C. Also included in this mixture to neutralize any potential VSV-G carryover virus was 1E9F9, a mouse anti-VSV Indiana G, at a concentration of 600 ng/ml (Cat#Ab01402-2.0, Absolute Antibody, Oxford, UK). The serum-virus mixture was then used to replace the media on VeroE6 TMPRSS2 cells. 23-24 hours post infection, the cells were washed and fixed with 4% paraformaldehyde before visualization on an S6 FluoroSpot Analyzer (CTL, Shaker Heights OH). Individual infected foci were enumerated and the values compared to control wells without antibody. The focus reduction neutralization titer 50% (FRNT_50_) was measured as the greatest serum dilution at which focus count was reduced by at least 50% relative to control cells that were infected with pseudo-type virus in the absence of patient serum. FRNT_50_ titers for each sample were measured in at least two technical replicates performed on separate days.

### Statistical analysis

Reciprocal serum dilution antibody titers were log2 transformed for statistical analysis. ELISA antibody titers below the limit of detection were set to a reciprocal titer of 25. Log2 transformed antibody titers were compared with one-way ANOVAs and unpaired t-tests. Statistical significance was set to p-value < 0.05. Linear regressions were also performed using log2 transformed titers and untransformed data from the other variables. Statistical analyses were performed using Prism version 8 (GraphPad Software, San Diego CA).

## Data Availability

All data are included in the manuscript.

## DATA AVAILABILITY

All data are included in the manuscript.

## ACKNOWLEDGEMENTS

EMA and TBM were supported by the NIH Training in Virology T32 Program through grant number T32-AI-007324. PH was supported by the NIH Emerging Infectious Diseases T32 Program T32AI055400. PB was supported by a Peer Reviewed Medical Research Program award PR182551 and grants from the NIH (R21AI129531 and R21AI142638). This work was supported by institutional funds from the University of Pennsylvania. We thank the COVID-19 Processing Unit (CPU) at the University of Pennsylvania for receiving and processing sera samples. We thank Jeffrey Lurie and we thank Joel Embiid, Josh Harris, David Blitzer for philanthropic support.

---

**Supplemental Table S1.** Comparative clinical features and laboratory data for each pediatric SARS-CoV-2 cohort. Data are presented as median [IQR]; *Included intubation with ventilation or non-invasive positive pressure ventilation; ^a^Co-infections included *Staphylococcus aureus* osteomyelitis (N = 2, with 1 of 2 also with bacteremia), Salmonella enteritis (N = 1); ^b^Co-infections included *E. coli* bacteremia (N = 1), Enterovirus meningitis (N = 1), Adenovirus, Rhinovirus, and calvarial osteomyelitis (N = 1), and *E.coli* urinary tract infection (N = 1); ^c^Coinfections included Parainfluenza virus infection (N = 1), possible Epstein-barr virus with positive IgM (N = 1) and Rhinovirus infection (N = 1); NT - not tested. Data were not available for all patients.

|  |  |  | Minimal<br>N=10 | Severe<br>N=9 | MISC-C<br>N=10 |
| --- | --- | --- | --- | --- | --- |
| Age in years [IQR] |  |  | 14 [3.5-16.5] | 16 [14-17] | 8.5 [6-14] |
| Presence of Co-infection, N (%) |  |  | 3 (38)<sup>a</sup> | 4 (44)<sup>b</sup> | 3 (30)<sup>c</sup> |
| ICU Admission, N (%) |  |  | 1 (12) | 9 (100) | 7 (70) |
| Respiratory support*, N (%) |  |  | 0 (0) | 9 (100) | 4 (40) |
| Inotropic support, N, (%) |  |  | 0 (0) | 7 (78) | 7 (70) |
| SARS-CoV-2 RT-PCR (Ct) |  |  | 34 [22-40] | 28 [26-29] | 38 [35-40] |
| SARS-CoV-2<br>antibody titers<br>(reciprocal<br>dilution) | S-RBD | IgG | 91 [$<50$-3624] | $<50$ [$<50$-304] | 3532 [982-8677] |
| | | IgM | 129 [$<50$-433] | $<50$ [$<50$-424] | 140 [$<50$-389] |
| | | IgA | $<50$ [$<50$-1026] | $<50$ [$<50$-55] | 69 [$<50$-3590] |
| | S | IgG | 227 [$<50$-9716] | $<50$ [$<50$-1039] | 7982 [1862-19597] |
| | | IgM | 113 [$<50$-650] | 166 [$<50$-602] | 138 [$<50$-307] |
| | | IgA | 163 [$<50$-2106] | $<50$ [$<50$-135] | 3015 [240-6814] |
| | N | IgG | 251 [139-1853] | $<50$ [$<50$-4843] | 2140 [604-3713] |
| | | IgM | $<50$ [$<50$-$<50$] | $<50$ [$<50$-573] | $<50$ [$<50$-$<50$] |
| | | IgA | $<50$ [$<50$-101] | $<50$ [$<50$-$<50$] | $<50$ [$<50$-124.5] |
| Ferritin |  |  | NT | 677 [181-9860] | 804 [686-892] |
| D-dimer |  |  | NT | 2.5 [0.8-20.5] | 5.8 [3.5-20.5] |
| C-reactive protein |  |  | 18.3 [14.6-25.5] | 30.9 [7.0-34.9] | 21.7 [19.1-33.0] |
| ESR |  |  | 51 [42-93] | 20 [13-33] | 68 [40-82] |
| BNP |  |  | NT | 395 [46.8-671] | 1003 [377-1554] |

